# Contribution of age-related effects to COVID-19 differences of crude fatality ratios in two Argentine provinces between March and August 2020

**DOI:** 10.1101/2020.11.28.20240127

**Authors:** Octavio Nicolas Bramajo, María Florencia Bathory

## Abstract

The following paper presents temporary estimations of CFR (the ratio between deaths and infected positive cases) attributed to COVID-19 for two provinces in Argentina (Jujuy and Buenos Aires Province), using public data provided by the Argentine Ministry of Health. In order to make comparisons between jurisdictions, we applied a series of exploratory measures (which resulted in excluding many other jurisdictions from the comparison), and later on the Kitagawa decomposition procedure, trying to separate rate (“net” fatality) and structure components (age-attributable effects) from CFR estimations in those provinces. After the decomposition we can observe that between almost non existant differences on average, the magnitude of structure and net rate effects tend to go into different directions across age groups, indicating some premature mortality in Buenos Aires and an excess net CFR in Jujuy for older age-groups

## Introduction

The pandemic caused by Coronavirus disease or COVID-19 has arguably been one of the most relevant events worldwide in the last hundred years. Up to this day. different efforts have been made in various disciplines to have a better understanding of the scope of the pandemic and its effects, not only on the health of populations, but in different areas of society, such as economics, education, leisure, among many others (and arguably all of them). Because the process it is still ongoing, many of the efforts to glimpse the magnitude of the consequences of the pandemic are transient in nature and should not be thought of as complete processes. In Argentina, the first case identified as positive by COVID-19 was on March 3, 2020. By the end of August of the same year, more than 400,000 positive cases detected and nearly 8,000 deaths have been counted, although this process has accelerated significantly in recent months along with the population’s testing capacity: as of June 1, nearly 16,800 cumulative positive cases and a total of 539 deaths were reported. These numbers may be artificially low on the impact of the disease given the difficulties of conducting massive tests in the countries of the region (1), but this work does not seek to question “the right number of cases” (although it is a relevant question in any estimation on the matter) in the country but to provide a descriptive and approximate picture of the CFR attributed to the pandemic.

Traditionally, age has been one of the determining factors when considering the negative health effects of most diseases. The case of COVID-19 is no exception, as particular emphasis has been placed on age as the main determinant of the risk of death caused by the disease, and in men as the main affected. This is because the observed CFR of the virus has mostly been concentrated in adult men over 65 years of age (2). In both Argentina and other Latin American countries, there are populations with a more rejuvenated age structure when compared to Europe, but with a higher prevalence of noncommunicable diseases caused by its own unequal epidemiological transition process (3), coupled with greater social inequality (4), more precarious housing conditions, less social protection, among other unfavorable structural situations that made the region particularly vulnerable to disease and its consequences (1.5). As is the case for other countries, in Argentina the demographic and epidemiological transition recorded different durations and sequences according to socio-economic sectors, in the urban and rural spheres, as well as in the geographical regions that make up the national territory (6). The early beginning of Argentina’s demographic transition was associated with the growth of the Pampas region and particularly in the vicinity of the region of what now makes up the space of Greater Buenos Aires (comprising the entire Autonomous City of Buenos Aires and a part of the territory of the Province of Buenos Aires, creating one of the largest Metropolitan Regions in Latin America, with about 20 million inhabitants). The results of this process are expressed that today the Autonomous City of Buenos Aires has the oldest population pyramid and one of the highest life expectancies in the country. And the provinces of the North and Northeast of the country the youngest population structures, being the regions that have presented a later social development in the aforementioned transitions (7.8). It is then to be expected that these various structural effects could affect subnational estimates of the net CFR caused by COVID-19. In other words, it is possible that some or all of the fatality estimates caused by the disease in Argentina may be hiding effects related to the age structure of the affected population (9), either by hiding or exaggerating some differentials presented to date.

### Objective

This work had the objective, on the one hand, to present an estimate of the case fatality ratio caused by COVID-19 in Argentina from its arrival in the country until the end of August 2020, both nationally and subnationally, through the separation of effects linked to the age structure of cases recorded through mathematical decomposition techniques. These inputs sought to provide a clearer picture of the fatality ratio of the virus at the general level, but also by identifying at which ages the differences are greater between provinces, detecting possible foci of premature mortality.

## Materials and Methods

An ecological type study was conducted from aggregated data to conduct a first exploration of differentials at the population level, performing some exploratory analysis in the first part of the study, and mathematical decomposition procedures on the second part. To do this, we took advantage of the daily report of data provided by the Ministry of Health of the Nation of Argentina (or MSAL, given their initials in Spanish), with information collected online through the Integrated Health Information System (SISA). In the case of anonymous, open and public data it was not necessary to obtain any informed consent or approval of any ethics committee for analysis. The report used to date was the count until August 31, 2020 (9). In these records it was possible to observe a number of basic sociodemographic characteristics about those who have been reported as suspected cases of COVID-19, and when the disease has been detected for reported positive cases. In addition, it is also recorded whether that person has died, and the date of death. However, it should be noted that such information may contain errors a resulting from manual loading of data, as well as ex post verifications and corrections may have been carried out by the health authorities. That is, while these data are constantly updated, these are not those provided and harmonized by the Office of Health Statistics and Information (DEIS), a bureau that deals with Argentina’s yearbooks of vital statistics, and should therefore be taken as provisional in nature. On the other hand, not every death that corresponds to a positive case implies that the cause of death was COVID-19. That is why emphasis is placed on the provisional nature of this study and the estimates obtained. It is not the object of this analysis to discern the “true” impact of the total CFR caused by COVID-19 on the Argentine population, but to make an approximation based on what is known to date and with the available sources. That being said, it also has to be noticed that case-detection (testing) is a very important aspect of this work: sometimes differences in CFR may result from differences in socio-sanitary conditions, but is also probable that those differentials are due to testing capacities. While we cannot establish clearly the cause of a given CFR difference between two jurisdictions, we can assume that provinces that have larger differentials in positive cases imply that is more likely that those differences are a result of undertesting, and that similar percentages of positive cases on the overall population may imply that the provinces are suitable for comparison. Therefore, part of the exploratory analysis will consist in visualizing the proportion of positive cases detected in the overall population.

So far, the case fatality ratio (CFR) has traditionally been presented in epidemiological analyses as an indicator of the strength of mortality of the pandemic, understood as the relationship between deaths (D) and positive cases (C) of the virus.

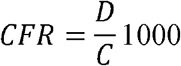

Where information on deaths distributed by age group is available, the CFR can also be expressed as the weighted sum of the different proportions of death (P) in the different five-year age groups (e):

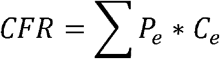

However, it is well known fact that dissimilar structures of the affected population can affect phenomena such as mortality and CFR and can mask and confuse the magnitude of the effects in the results obtained. Typically, standardized rates are used for this type of analysis. But the technique has an important limitation: the results are expressed according to an arbitrary standard that does not allow to see the net effects of the phenomenon in question. So instead the technique of decomposition of effects suggested by Kitagawa (10) This procedure (also known as Oaxaca-Blinder Decomposition) serves to separate, in a difference of two rates corresponding to two groups G1 and G2, how much of the difference can be explained by the net effect corresponding to the incidence of the phenomenon in question (also known as “Rate Effect”, or in this case “RE”) and how much of that difference responds to a compositional effect (attributable to the age structure of the groups, known as “Composition Effect” or “Structure Effect”, or “SE” in this case).

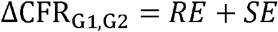

This method is very useful for breaking down effects between groups/populations for which only one observation is taken at a given time, and above all it has already been used satisfactorily to analyze the differences in CFR caused by COVID-19 in other countries (11). In addition, like general decomposition methods, it allows to disaggregate the different effects in age groups (ten age groups in this case, considering the available cases and deaths), identifying groups where these differences expand and contract, allowing to identify potentially vulnerable groups where the “net” CFR is greater. After quality checks, a minimum number of cases (less than 1%) were noticed. for which the province or sex was not registered. The first (corresponding to 8 deaths) have been excluded from the fee analysis, while the latter (53 deaths distributed almost entirely in the Autonomous City of Buenos Aires and the Province) were imputed at different ages and sexes with a simple criterion of proportionality (12). To simplify the analysis, only seven jurisdictions were considered for the exploratory analysis: Province of Buenos Aires, Autonomous City of Buenos Aires, Chaco, La Rioja, Jujuy, Río Negro and Mendoza, which are the ones with the greatest CFR since the remaining ones do not present enough cases to allow their inclusion at such a level of disaggregation. Similarly, only the mortality of individuals between the age of 30 and 99 were only considered to avoid excessive weights of the low CFR of the virus in children and young people and to avoid potential “noises” with centenaries. It is worth mentioning the socio-economic and health heterogeneity that the selected jurisdictions present: on the one hand Chaco, La Rioja and Jujuy are three jurisdictions that are in the lower half of the distribution of The Geographical Gross Product per Capita in Argentina, as opposed to the remaining four (8). Likewise, it is emphasized that Chaco and Jujuy are located in the lower quartile of life expectancy at birth in Argentina, while in the upper quartile are located the Autonomous City of Buenos Aires, Río Negro and Mendoza, three of the most feminized jurisdictions in the country (8). Therefore, it is also worth considering these heterogeneities and disparities at the regional level when considering the results of this work.

## Results

### Exploratory Analysis

Table 1 presents the distribution by jurisdiction of cases and deaths among individuals 30 and 99 years of age. According to population projections by the National Institute of Statistics and Census (INDEC), the Province of Buenos Aires and the Autonomous City of Buenos Aires (CABA) present 45% of the population of Argentina as of 2020 (13), so it is not surprising that they are the jurisdictions that present the greatest number both of cases and deaths, followed by Jujuy, Chaco, Río Negro, Mendoza and La Rioja respectively. 55.7% of total deaths in all 7 jurisdictions are male, and in all of them this indicator exceeds 50%. Only in CABA some parity in deaths could be considered given the data.

**Table 1:** Distribution of deaths recorded by jurisdiction as of August 31, 2020.

| Province | Cases | Deaths | % Male Deaths |
| --- | --- | --- | --- |
| Buenos Aires | 264681 | 8454 | 56,2 |
| CABA | 97917 | 3040 | 52,2 |
| Jujuy | 9005 | 327 | 64,1 |
| Chaco | 5502 | 217 | 62,7 |
| Río Negro | 5876 | 182 | 62,5 |
| Mendoza | 7419 | 163 | 64,4 |
| La Rioja | 2088 | 80 | 59,9 |
| **Total** | **277090** | **12463** | **55,7** |

Source: authors' calculations based on SISA/MSAL (10)

**Table 2:** CFR (by 1000) in ages 30-99 and results of Kitagawa decomposition for COVID-19 CFR in Buenos Aires and Jujuy provinces, March-August 2020

| Province | **CFR**<br>**(\*1000)** | ΔCFR<br>*1000 | RE*1000 | SE*1000 | RE<br>(relative) | SE<br>(relative) |
| --- | --- | --- | --- | --- | --- | --- |
| Buenos Aires | 45.00 | - | - | - | - | - |
| Jujuy | 44.95 | 0.05 | 0.33 | -0.28 | 54% | 46% |

Source: authors' calculations based on SISA/MSAL (10)

Before delving deeper into the results decomposition, Figure 2 provides the proportion of positive cases by province, and we can confirm that Chaco, the province with the highest CFR, has also one of the lowest proportions of positive cases detected (which could suggest an important lack of detection tests). And CABA, the capital city, has, by large, the highest proportion of positive cases, which means that the rates may also be misleading when comparing with the other provinces (because any decomposition will overestimate both rate effects and the age-structure effect). After visual inspection, we conclude that the most feasible comparison for a decomposition analysis should be between the Province of Buenos Aires and Jujuy.

**Figure 1:**
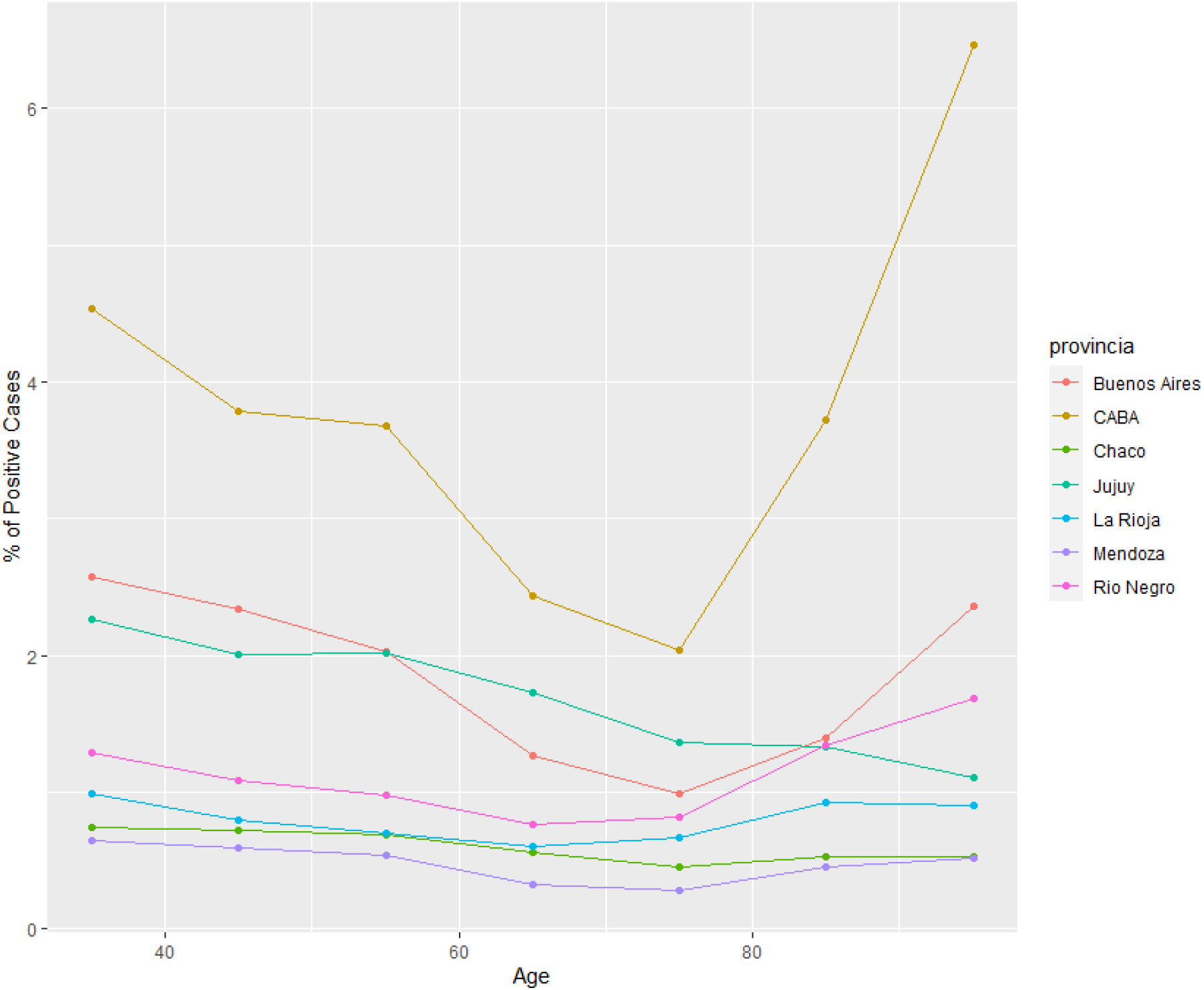
COVID-19 percentage of positive cases by sex and ten age groups in given Argentine provinces, March-August 2020 <u>Source:</u> authors’calculations based on SISA/MSAL (10)

**Figure 2:**
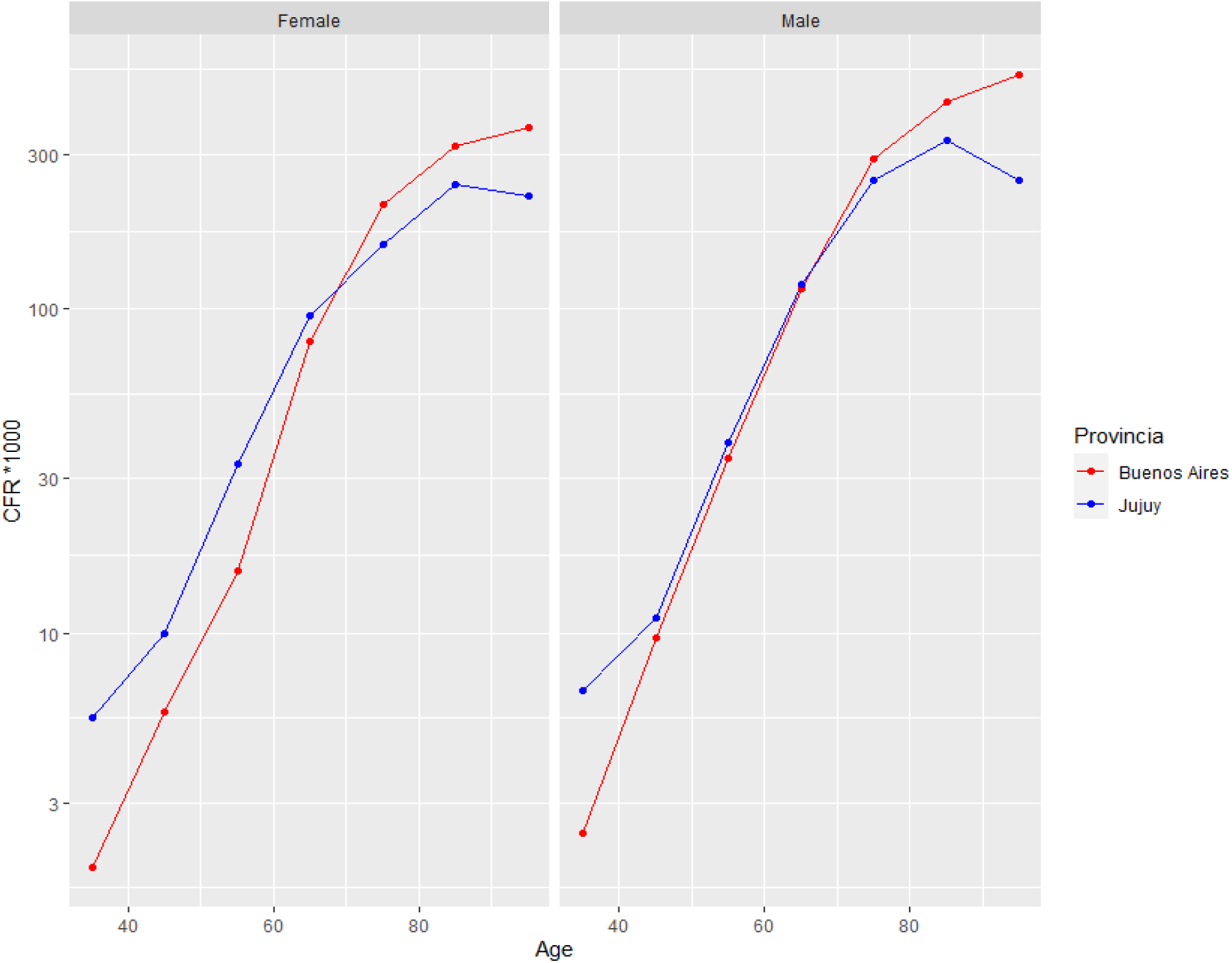
COVID-19 CFR (by thousand) by sex and ten age groups in Buenos Aires y Jujuy Provincias al 31 de Agosto 2020 <u>Source:</u> authors’calculations based on SISA/MSAL (10)

Figure 2 indicates the evolution of CFR recorded by sex and age in the two selected provinces, that apparently tend to show similar patterns. It is observed, on the one hand, that CFR grows exponentially with age, slowing only at advanced ages, past 80 years. On the other hand, it is confirmed that in the age groups analyzed the male CFR is greater than CFR for females.

### Decomposition Results

Table presents the results of Kitagawa’s decomposition. We are using Buenos Aires as the Province that serves as a reference for comparisons with Chaco (remember that differences in CFR rates are expressed as “ΔCFR” and “SE+ RE”). The results indicate that while the overall CFR for both provinces is practically the same, there is slight difference by structure: if both provinces would have the same age structure between positive cases, the difference between Buenos Aires and Jujuy would be slightly larger (favoring the former), and age-structure components explain almost half of the observed CFR difference, albeit it is small in their magnitude.

As mentioned above, it is also possible to visualize the different contribution of components by different age groups, as presented in Figure 3, again using Buenos Aires as a reference. We can see an important difference of effect distribution, that end up compensating each other: After age 70 it becomes clear that there is a strong age-structure effect that diminishes the observed CFR difference, but before that point, it seems the other way round (which could indicate some undercounting of cases in Buenos Aires province). Furthermore, after age 80 there is an important net difference in CFR that indicates that differentials should be larger than the observed, if both provinces had the same exposure.

**Figure 3:**
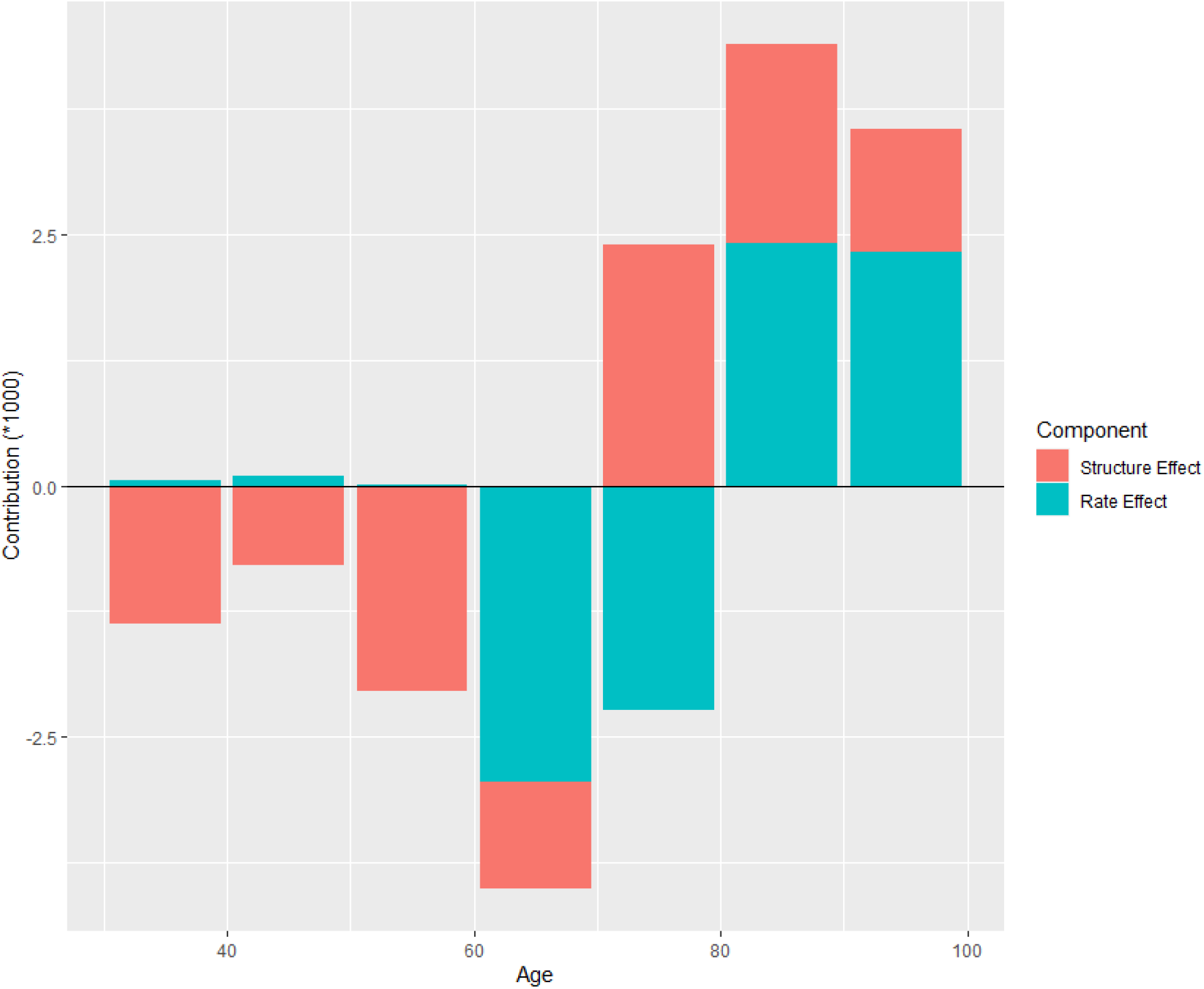
Contribution of components by age between Buenos Aires and Jujuy in CFR differences by COVID-19, March-August 2020 <u>Source:</u> authors’calculations based on SISA/MSAL (10)

## Conclusions

This work aimed to illustrate a descriptive and comparative picture of mortality from COVID-19 in Argentina, emphasizing the jurisdictions that were most affected by the pandemic between march and August. After some exploratory analysis, regrettably the case-detection capacities in the different provinces make most of them unsuitable for analysis. However, a comparison is possible between the Buenos Aires province (the largest of the country in population size) and the northern province of Jujuy. We can confirm that in both provinces CFR and testing patterns seem similar, that that CFR grows exponentially with age, and that CFR is higher for males than females. Furthermore, the decomposition analysis shows that differences between the jurisdictions, albeit small in magnitude, would be slightly larger if both provinces would have the same age-structure in positive cases. By presenting component differences in age groups for different jurisdictions, it is appreciated that the small magnitude differences in CFR are actually a result of components going into opposite directions that in average tend up nullifying themselves. This would suggest higher premature mortality in Buenos Aires, but a higher net CFR in Jujuy, along with differences in the compositional effects.

## Limitations

It is also worth mentioning the limitations presented by this work: it is worth remembering that there were reasons in the scientific and health community to think that Latin America could be one of the regions most hit by the pandemic caused by COVID-19 (Nepomuceno et al., 2020). While this is still a fledgling phenomenon, Argentina appears to be no exception: to date there are no signs indicating a slowdown in mortality that the virus could cause. Therefore, the estimates presented here are of a partial type, which must be taken into account when analyzing them. Second, the CFR is a simple but inaccurate indicator of attributable mortality in a population. While some of these effects can be corrected with mathematical procedures (decompositions, standardizations), there are other situations that the indicator cannot account for: mainly asymptomatic infected ones, which are not recorded as positive cases because they have not been tested. While the fatality rate may account for early trends in a pandemic, perhaps in the long run other indicators will be more sophisticated (11.14). There may also be deaths attributed to COVID-19 that actually correspond to other causes and vice versa, as well as other omissions delays in burdens. It is also worth remembering that we are working here with registration instruments that are not intended for demographic analysis. Therefore, once vital statistics data becomes available, parsing with the information presented would be desirable. In addition, the distribution of positive cases by age itself may be affected by the different age structures of the population (something for which it is not controlled in this work). On the other hand, the small number of cases, while allowing an overall analysis of the components of CFR, does not allow detailed distinctions to be made by sex when breaking down the effects caused by differences in fatality rates.

## Discussion

Despite these limitations, this work succeeded in establishing that the effects attributable to the age structure only explains a portion of the COVID-19 CFR differences established between two jurisdictions in Argentina, identified in which age groups these differences are greater and lower, and illustrated a simple mathematical decomposition procedure to make the different rates of CFR in two provinces of the country.

## Data Availability

The data is available freely upon request to the authors

